# ChatGPT vs DeepSeek: A Comparative Study of Diagnostic Accuracy and Clinical Reasoning in Rare and Complex Diseases

**DOI:** 10.1101/2025.08.28.25331796

**Authors:** Jialin Liu, Weiping Cao, Bo Yuan, Wenyi Xie, Changyu Wang, Siru Liu

## Abstract

Diagnostic errors in rare and complex diseases contribute significantly to morbidity and mortality. The ability of large language models (LLMs) to enhance diagnostic performance in such cases remains uncertain. This study compares the diagnostic accuracy, clinical reasoning quality, and inference efficiency of three ChatGPT variants (o3-mini, o3-mini-high, o1) and DeepSeek-R1 using 30 English-language case reports of rare and complex diseases from 26 specialties across 15 countries, sourced from PubMed and Web of Science Core Collection databases. Cases were selected to avoid overlap with model training data. Each case was processed once by each model, with outputs anonymized and evaluated in a double-blind manner by two board-certified physicians (each with >15 years’ clinical experience) and ChatGPT-4o. Diagnostic accuracy, the primary outcome, ranged between 30.0% and 40.0% with no significant differences observed among models (Cochran’s Q test, P = 0.16). ChatGPT-o1 achieved the highest accuracy (12/30, 40.0%; 95% CI, 24.6%+/-57.7%), followed by ChatGPT-o3-mini and o3-mini-high (each 11/30, 36.7%), and DeepSeek-R1 (9/30, 30.0% for each English and Chinese language inputs). Mean reasoning scores differed significantly (P < 0.05): ChatGPT-o1, 4.08 +/- 0.82; DeepSeek-R1 (English), 3.86 +/- 0.86; ChatGPT-o3-mini, 3.71 +/- 0.90; ChatGPT-o3-mini-high, 3.69 +/- 0.80; DeepSeek-R1 (Chinese), 3.67 +/- 0.84. Inter-evaluator agreement was high (ICC = 0.84; 95% CI, 0.80-0.88). Inference times varied significantly (P < 0.001), with ChatGPT-o3-mini being fastest (7.0 +/- 3.8 s) and DeepSeek-R1 (English) slowest (46.5 +/- 32.5 s). Advanced LLMs demonstrate potential to support diagnosis of rare and complex diseases, with transparent reasoning processes that may aid clinical decision-making and medical education. Further domain-specific refinement and prospective clinical validation are essential for safe and effective integration into clinical practice.

**Highlights:**

- While LLMs showed similar diagnostic accuracy (30-40%) in rare and complex diseases, ChatGPT-o1 significantly excelled in the quality of its clinical reasoning.
- Inference speeds varied dramatically (7s-47s), highlighting a critical trade-off between model performance and real-world utility.
- The transparent reasoning of LLMs shows clear promise as a tool to support clinical decision-making and medical education.
- Safe clinical implementation is dependent on future domain-specific refinement and prospective validation.

## Introduction

Diagnostic errors present a significant challenge in clinical practice, particularly with regard to rare and complex diseases, and substantially contribute to morbidity and mortality rates.^1,2^ In U.S. outpatient settings, diagnostic errors affect approximately 5% of adults,^3^ while among hospitalized patients, they account for 6% to 17% of adverse events,^4^ compromising patient safety and healthcare quality. Annually, these errors result in severe harm to an estimated 795,000 patients in the U.S. (range, 598,000–1,023,000), including approximately 371,000 fatalities and 424,000 cases of permanent disability.^5^ Rare and complex diseases are particularly challenging due to atypical presentations, limited diagnostic resources, and the need for specialized expertise, often leading to prolonged diagnostic delays.^6^ Clinicians face high workloads, cognitive demands, and increasing burnout rates, which exacerbate cognitive biases and reduce diagnostic accuracy.^7^

Advances in artificial intelligence, particularly large language models (LLMs), offer a promising approach to mitigate these challenges by supporting clinical reasoning and decision-making.^8^ While LLMs have shown efficacy in medical knowledge assessments and diagnosing common diseases,^8–11^ their application to rare and complex diseases remains emerging, with limited evidence on their diagnostic accuracy and clinical utility.^12–14^ This study evaluates the diagnostic performance and reasoning quality of four LLMs—ChatGPT-o3-mini, ChatGPT-o3-mini-high, ChatGPT-o1, and DeepSeek-R1—using real-world cases of rare and complex diseases. By comparing their diagnostic accuracy, reasoning processes, and inference efficiency against clinical expert standards, we aim to elucidate their potential to enhance diagnostic decision-making and inform the development of advanced diagnostic support systems.

## Methods

### Case selection

We selected 30 clinical case reports of rare and complex diseases from PubMed and Web of Science Core Collection databases, published between January 1, 2024, and February 15, 2025. The search strategy used the keywords “rare disease” OR “complex disease” AND “case report” (title). Records indexed through February 15, 2025, were included, which postdate the December 31, 2024, cutoff for the training data of the large language models (ChatGPT-o3-mini, ChatGPT-o3-mini-high, ChatGPT-o1, DeepSeek-R1). From 806 initial records, duplicates were removed, and two independent reviewers screened titles and abstracts using predefined eligibility criteria. Full texts were assessed for: (1) publication in English in a peer-reviewed journal; (2) comprehensive clinical data (patient history, physical examination, laboratory results, treatment course); (3) detailed narrative descriptions of imaging or multimedia content sufficient for text-based review; (4) representation of diverse clinical specialties (e.g., internal medicine, neurology, oncology, surgery, pediatrics), with the 30 cases collectively spanning at least 26 diverse clinical specialties; (5) balanced geographic representation (Asia, Africa, Europe, North America, South America, Oceania), age groups, sex, and ethnicity; (6) explicit documentation of diagnostic reasoning; and (7) confirmation by a multidisciplinary panel of three board-certified clinical experts. Exclusion criteria included duplicate reports, absence of a definitive diagnosis, or incomplete clinical reasoning descriptions. Disagreements were resolved by consensus or adjudicated by a third expert using a four-point scale (1 = strongly exclude, 4 = strongly include), with cases scoring ≥3 by all experts included. Stratified random sampling ensured balanced representation across specialties and demographics.

### Models application

The study, conducted in February 2025, evaluated four LLMs: ChatGPT-o3-mini, ChatGPT-o3-mini-high, ChatGPT-o1, and DeepSeek-R1. Each model processed the 30 cases once using the prompt: “What is the most likely diagnosis?” Internet access was disabled to prevent external information retrieval. Inputs for ChatGPT models were in English, while DeepSeek-R1 received inputs in English and Chinese (translated by two bilingual experts and validated by ChatGPT-4o for semantic equivalence). Models used default configurations, employing chain-of-thought (CoT) prompting without additional fine-tuning, reflecting typical usage scenarios.

### Reasoning analysis rating scale

To assess the quality of clinical reasoning, five specialists from neurology, geriatrics, otolaryngology, oncology, and nephrology collaboratively developed a 5-point rating scale. The scale was grounded in established principles of diagnostic reasoning and designed to evaluate three key domains: evidentiary completeness, logical coherence, and clinical relevance.^15–17^ Reasoning was rated on a scale from 1 to 5. 1. Reasoning is entirely unclear, with no logical thread. 2. Poorly structured reasoning and scattered evidence. 3. Broadly complete reasoning but with noticeable logical gaps or insufficient support. 4. Logical, evidence-based reasoning that correctly includes the diagnosis within the differential, albeit with minor omissions.5. Complete, rigorously logical reasoning consistent with medical consensus, yielding a correct and well-justified diagnosis.

### Evaluation of model reasoning and discrepancy resolution

To evaluate LLM-generated clinical reasoning, we conducted a double-blind assessment using a panel of two board-certified physicians (each with over 15 years of experience in different specialties) and ChatGPT-4o. Evaluators underwent standardized rubric-based training, including calibration exercises, to ensure rating consistency. Model outputs were anonymized and randomly ordered. Each evaluator received the clinical case narrative, reference diagnosis with rationale, and a validated 5-point Likert scale rubric assessing logical coherence, evidentiary completeness, and clinical relevance. Materials were provided in Chinese and English, with translations independently verified for accuracy. Cases with a ≥2-point score discrepancy between evaluators were flagged for structured discussion between the physicians. If consensus was not reached, a third blinded clinical expert provided final adjudication. Inter-evaluator reliability was measured using the intraclass correlation coefficient (ICC).

### Statistical analysis

All analyses were performed using Python 3.8 (pandas 1.2.5, SciPy 1.7.3, statsmodels 0.12.2). Diagnostic accuracy was reported as proportions with 95% confidence intervals (CIs). Cochran’s Q test evaluated overall differences in binary outcomes across models, with pairwise comparisons conducted using McNemar’s test and Holm’s Bonferroni correction. To assess the differences in continuous variables among the groups, one-way analysis of variance (ANOVA) was performed. Post hoc pairwise comparisons with Bonferroni correction were conducted to identify significant differences between specific groups. Inter-evaluator reliability was assessed using two-way random-effects ICC, interpreted as: 0.90 excellent.^18^ A two-sided P < 0.05 was considered statistically significant.

### Ethical approval

Given the study utilized publicly available and anonymized data previously reviewed by an ethics committee, further ethical approval was waived.

## Result

### Clinical cases

The 30 selected cases, sourced from 15 countries across six continents and published in 17 peer-reviewed journals, spanned 26 clinical specialties, including oncology, neurology, neurosurgery, infectious diseases, hematology, general surgery, pediatrics, gynecology, immunology, otolaryngology, ophthalmology, allergology, psychosomatic medicine, and oral and maxillofacial surgery. This diverse representation reflects the complexity of real-world diagnostic challenges in rare and complex diseases (Table 1).

**Table 1.**
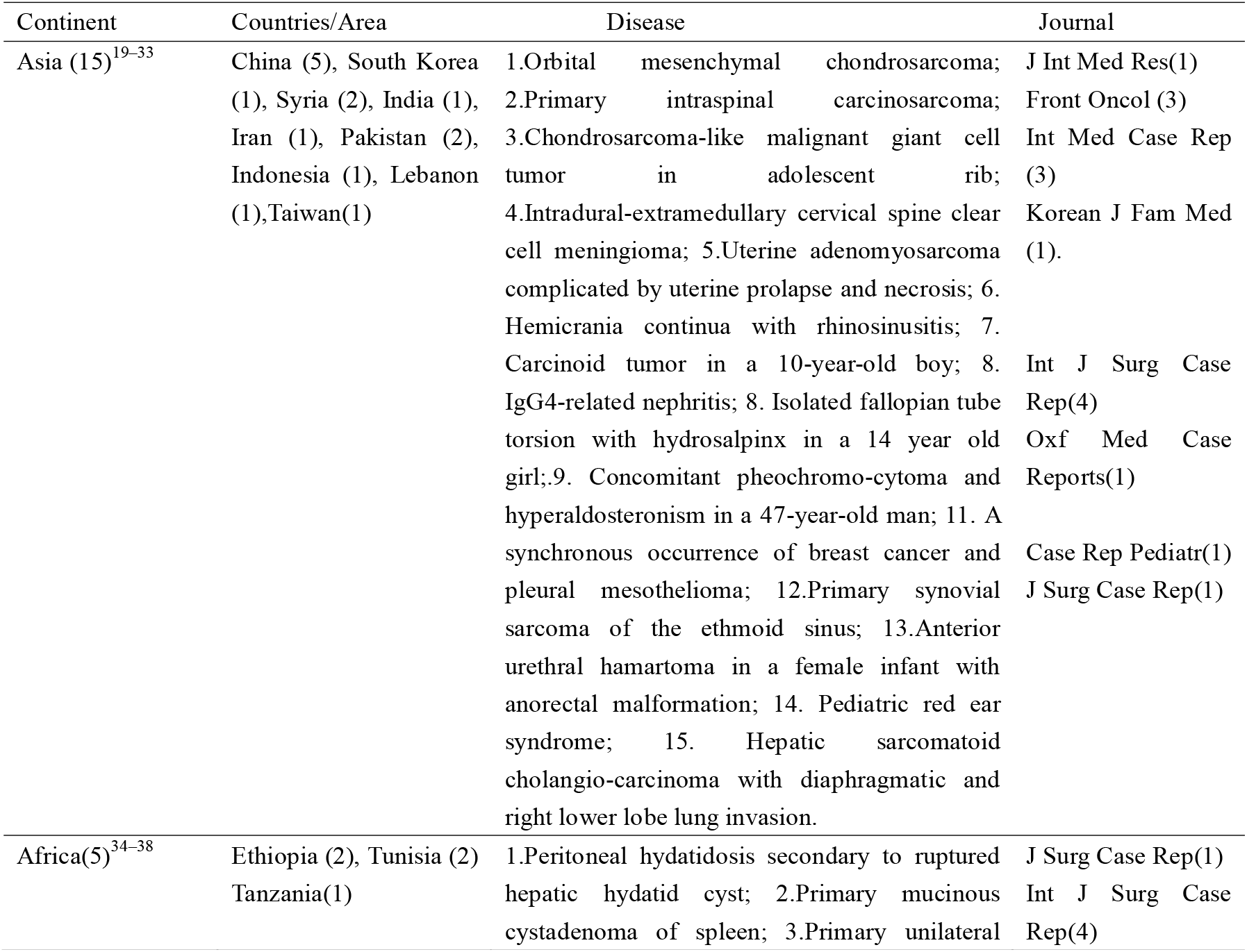
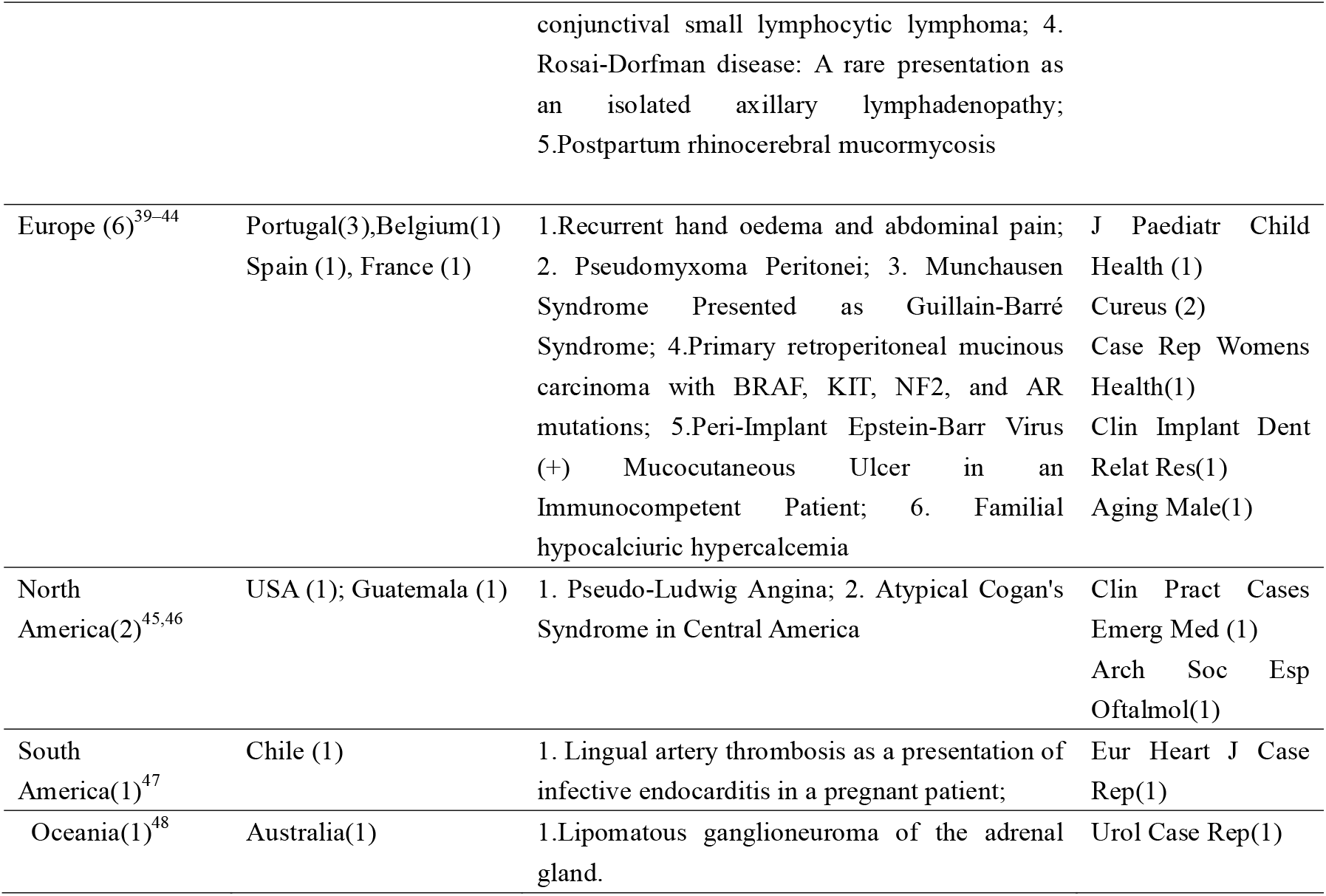
Geographic origin and publication sources of the 30 rare and difficult cases.

| Continent | Countries/Area | Disease | Journal |
| --- | --- | --- | --- |
| Asia (15)<sup>19–33</sup> | China (5), South Korea (1), Syria (2), India (1), Iran (1), Pakistan (2), Indonesia (1), Lebanon (1), Taiwan(1) | 1.Orbital mesenchymal chondrosarcoma; 2.Primary intraspinal carcinosarcoma; 3.Chondrosarcoma-like malignant giant cell tumor in adolescent rib; 4.Intradural-extramedullary cervical spine clear cell meningioma; 5.Uterine adenomyosarcoma complicated by uterine prolapse and necrosis; 6. Hemicrania continua with rhinosinusitis; 7. Carcinoid tumor in a 10-year-old boy; 8. IgG4-related nephritis; 8. Isolated fallopian tube torsion with hydrosalpinx in a 14 year old girl; 9. Concomitant pheochromo-cytoma and hyperaldosteronism in a 47-year-old man; 11. A synchronous occurrence of breast cancer and pleural mesothelioma; 12.Primary synovial sarcoma of the ethmoid sinus; 13.Anterior urethral hamartoma in a female infant with anorectal malformation; 14. Pediatric red ear syndrome; 15. Hepatic sarcomatoid cholangio-carcinoma with diaphragmatic and right lower lobe lung invasion. | J Int Med Res(1)<br>Front Oncol (3)<br>Int Med Case Rep (3)<br>Korean J Fam Med (1).<br>Int J Surg Case Rep(4)<br>Oxf Med Case Reports(1)<br>Case Rep Pediatr(1)<br>J Surg Case Rep(1) |
| Africa(5)<sup>34–38</sup> | Ethiopia (2), Tunisia (2) Tanzania(1) | 1.Peritoneal hydatidosis secondary to ruptured hepatic hydatid cyst; 2.Primary mucinous cystadenoma of spleen; 3.Primary unilateral conjunctival small lymphocytic lymphoma; 4. Rosai-Dorfman disease: A rare presentation as an isolated axillary lymphadenopathy; 5.Postpartum rhinocerebral mucormycosis | J Surg Case Rep(1)<br>Int J Surg Case Rep(4) |
| Europe (6)<sup>39-44</sup> | Portugal(3),Belgium(1)<br>Spain (1), France (1) | 1.Recurrent hand oedema and abdominal pain; 2. Pseudomyxoma Peritonei; 3. Munchausen Syndrome Presented as Guillain-Barré Syndrome; 4.Primary retroperitoneal mucinous carcinoma with BRAF, KIT, NF2, and AR mutations; 5.Peri-Implant Epstein-Barr Virus (+) Mucocutaneous Ulcer in an Immunocompetent Patient; 6. Familial hypocalciuric hypercalcemia | J Paediatr Child Health (1)<br>Cureus (2)<br>Case Rep Womens Health(1)<br>Clin Implant Dent Relat Res(1)<br>Aging Male(1) |
| North America(2)<sup>45,46</sup> | USA (1); Guatemala (1) | 1. Pseudo-Ludwig Angina; 2. Atypical Cogan's Syndrome in Central America | Clin Pract Cases Emerg Med (1)<br>Arch Soc Esp Oftalmol(1) |
| South America(1)<sup>47</sup> | Chile (1) | 1. Lingual artery thrombosis as a presentation of infective endocarditis in a pregnant patient; | Eur Heart J Case Rep(1) |
| Oceania(1)<sup>48</sup> | Australia(1) | 1.Lipomatous ganglioneuroma of the adrenal gland. | Urol Case Rep(1) |

### Diagnostic accuracy and inference time

Diagnostic accuracy ranged between 30.0% and 40.0% across models, with no statistically significant differences identified (Cochran’s Q test, P = 0.162). ChatGPT-o1 achieved the highest accuracy (12/30, 40.0%; 95% CI, 24.6%–57.7%), followed by ChatGPT-o3-mini and ChatGPT-o3-mini-high (each 11/30, 36.7%; 95% CI, 21.9%–54.5%), and DeepSeek-R1 (9/30, 30.0%; 95% CI, 16.7%–47.9% for both English and Chinese inputs). No significant differences were observed in pairwise comparisons (McNemar’s test, Holm-corrected P > 0.05) (Table2).

**Table 2.** Diagnostic accuracy and inference time of the LLMs.

|  | ChatGPT-o3-mini | ChatGPT-o3-mini-high | ChatGPT-o1 | DeepSeek-R1 English | DeepSeek-R1 Chines | p |
| --- | --- | --- | --- | --- | --- | --- |
| Accuracy | 36.7% | 36.7% | 40.0% | 30.0% | 30.0% | > .05 for all <sup>a</sup> |
| 95% CI | [21.9-54.5%] | [21.9-54.5%] | [24.6-57.7%] | [16.7-47.9%] | [16.7-47.9%] |  |
| Time (s) | 7.0±3.8 | 12.4±7.9 | 13.4±11.7 | 46.5±32.5 | 30.7±22.7 | > .05 for GPTo3-mini-high vs GPTo1<sup>b</sup><br>< .05 for others <sup>c</sup> |
| Rang | (3-17) | (3-41) | (3-49) | (16-161) | (11-137) |  |

a: Cochran p-value: 0.162; b: ChatGPT-o3-mini-high vs ChatGPT-o1:0.773; c: ChatGPT-o3-mini vs ChatGPT-o3-mini-high: <0.001 , ChatGPT-o3-mini vs ChatGPT-o1: <0.001, ChatGPT-o3-mini vs DeepSeek-R1 English: <0.001 , ChatGPT-o3-mini vs DeepSeek-R1 Chines: <0.001 ; ChatGPT-o3-mini-high vs DeepSeek-R1 English: <0.001 , ChatGPT-o3-mini-high vs DeepSeek-R1 Chines: <0.001 , DeepSeek-R1 English vs DeepSeek-R1 Chines:0.008.

Inference times differed significantly (P < 0.001). ChatGPT-o3-mini was fastest (7.0 ± 3.8 s; range 3–17 s), followed by ChatGPT-o3-mini-high (12.4 ± 7.9 s; range 3–41 s) and ChatGPT-o1 (13.4 ± 11.7 s; range 3–49 s). DeepSeek-R1 had the longest inference times (English: 46.5 ± 32.5 s, range 16–161 s; Chinese: 30.7 ± 22.7 s, range 11–137 s), both significantly longer than all other models (p < 0.05). Pairwise comparisons showed no significant difference between ChatGPT-o3-mini-high and ChatGPT-o1 (P = 0.773) (Figure 1, 2 and Table 2).

**Figure 1.**
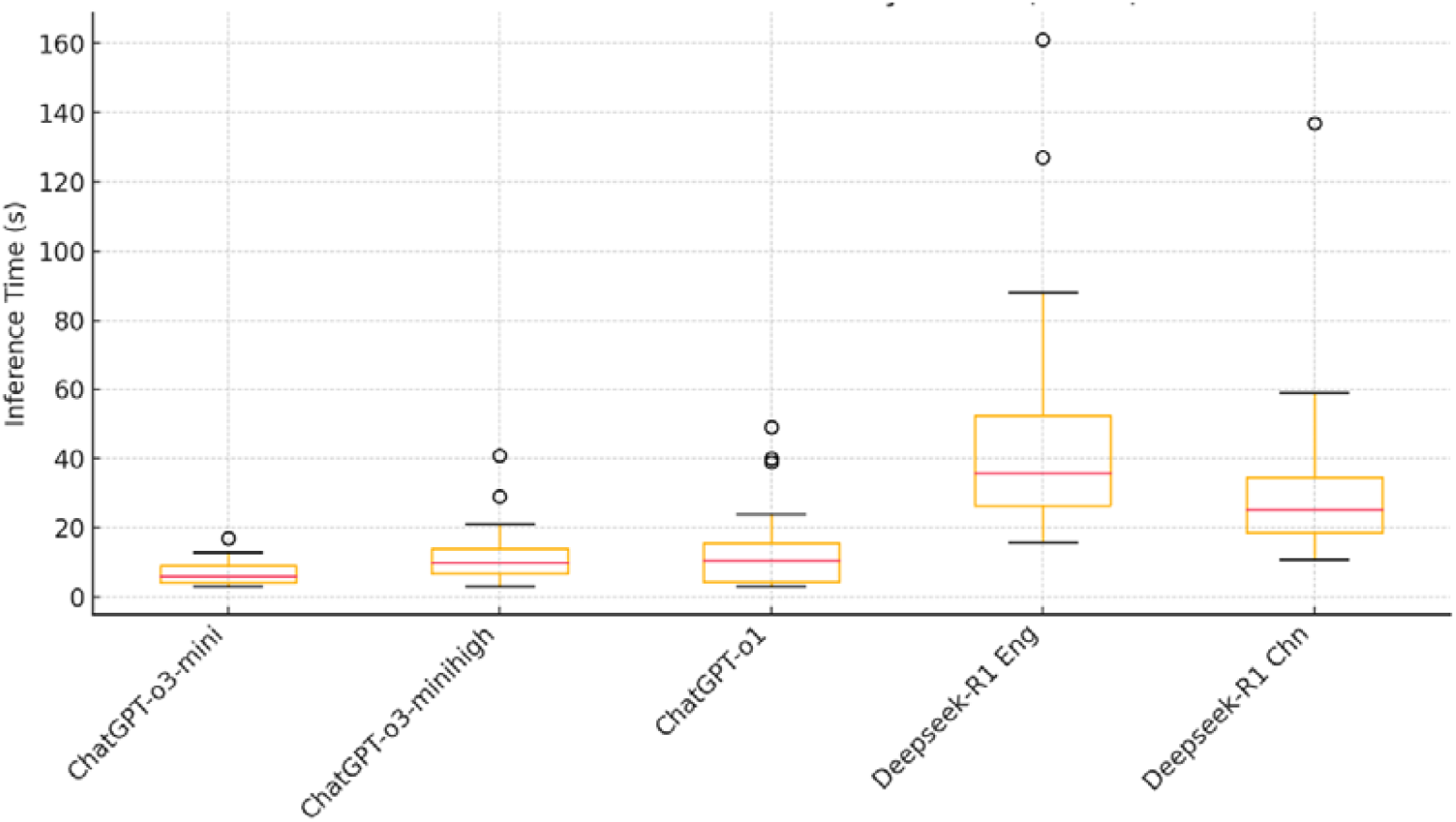
Inference time distribution by model (n=30)

**Figure 2.**
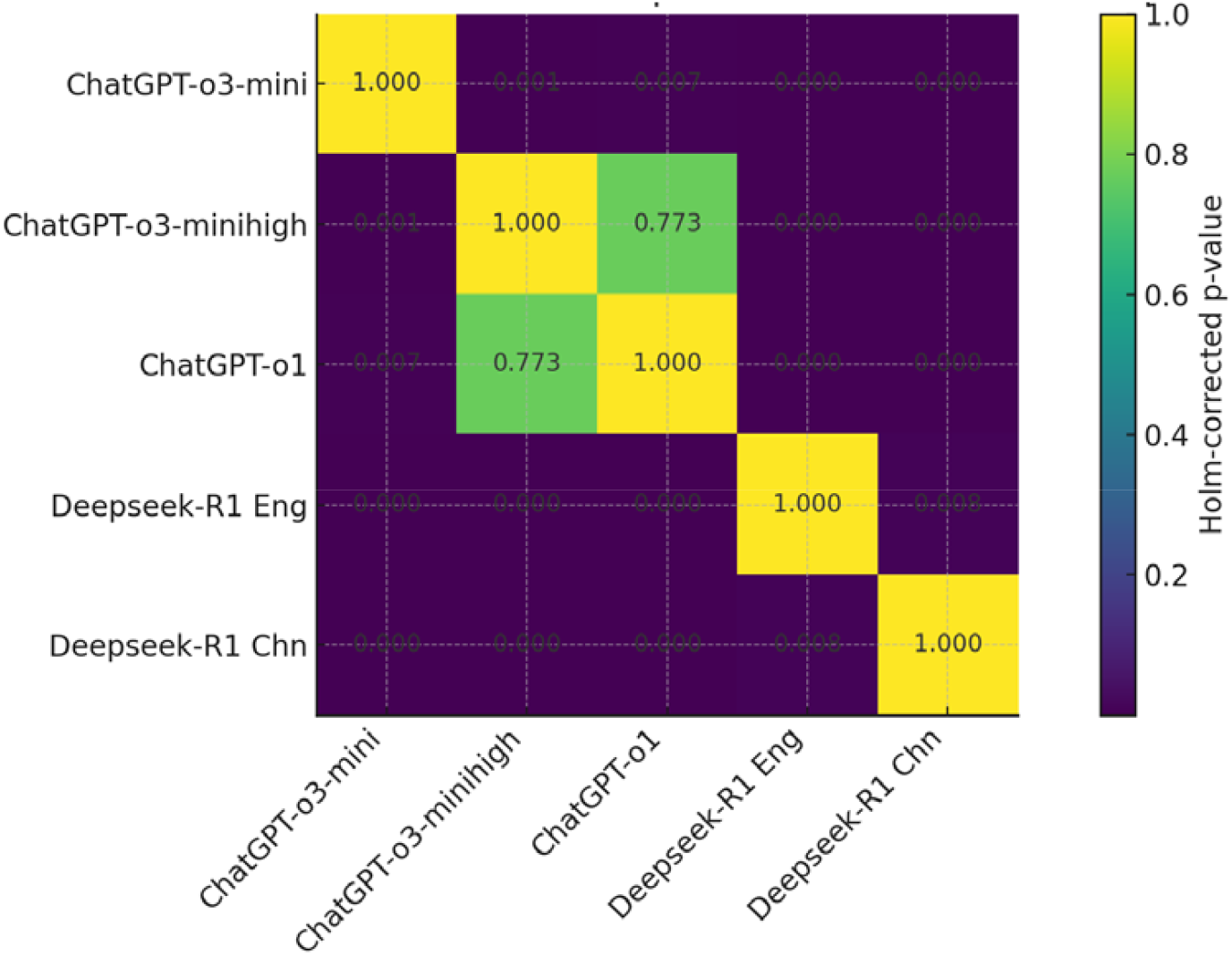
Heatmap of holm-corrected P-values for inference time

### Model Reasoning Evaluation

Reasoning scores showed high inter-evaluator agreement (ICC = 0.84; 95% CI, 0.80–0.88). ChatGPT-o1 achieved the highest mean reasoning score (4.08 ± 0.82), significantly outperforming other models (P < 0.05). DeepSeek-R1 (English) scored 3.86 ± 0.86, followed by ChatGPT-o3-mini (3.71 ± 0.90), ChatGPT-o3-mini-high (3.69 ± 0.80), and DeepSeek-R1 (Chinese) (3.67 ± 0.84). No significant differences were observed among the latter three models (P > 0.05) (Table 3 and Figure 3).

**Table 3.**
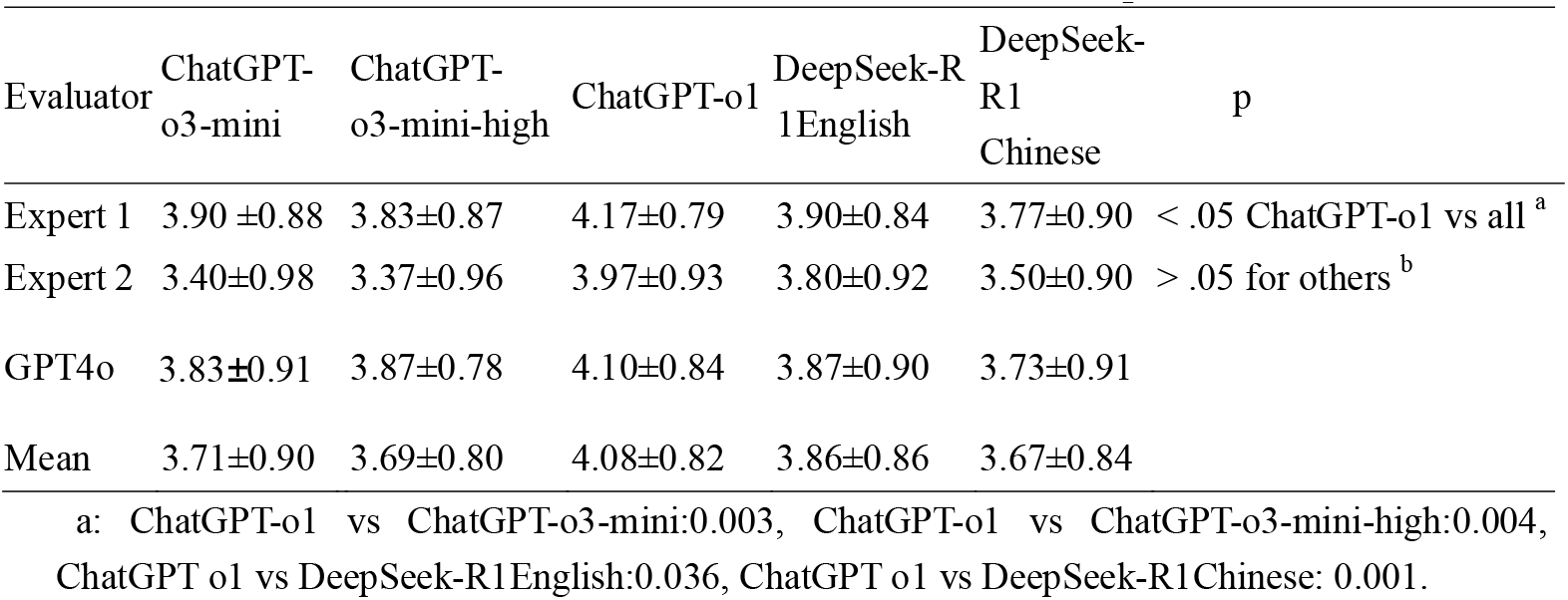
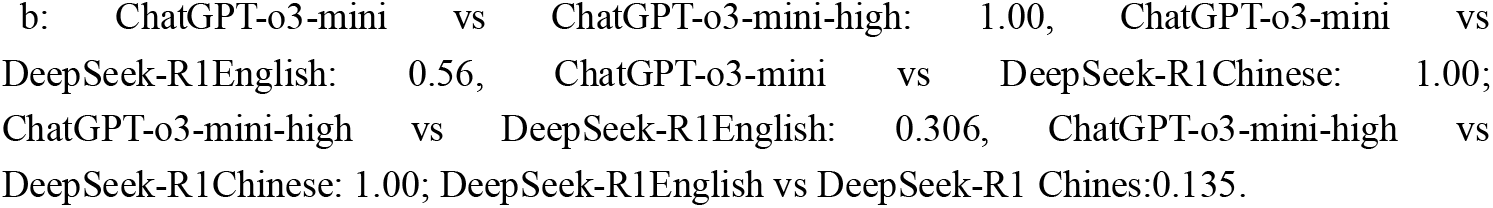
Inference Scores for ChatGPT and DeepSeek-R1 Models.

**Figure 3.**
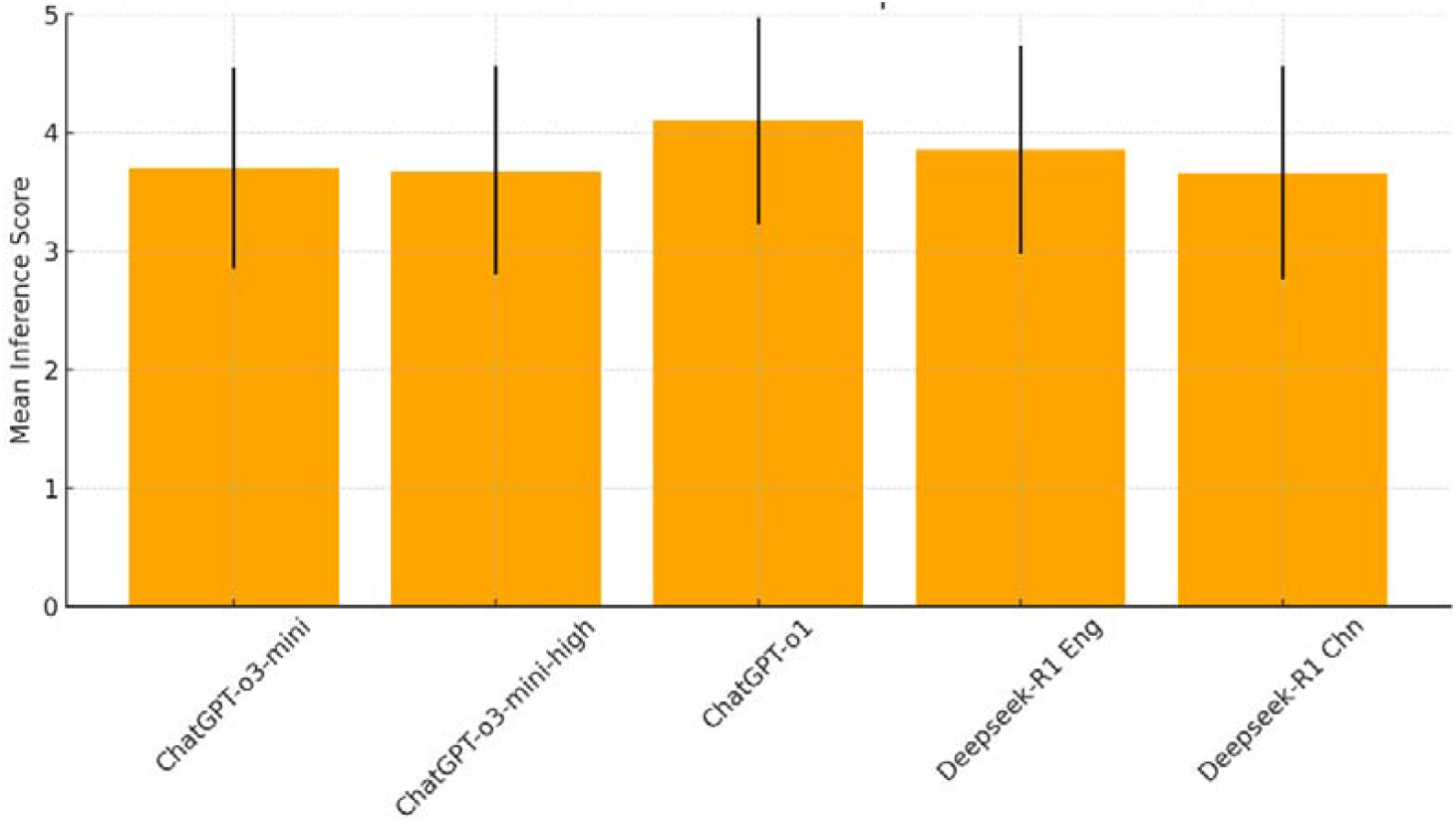
Mean Inference Scores for ChatGPT and DeepSeek-R1 Models (n=30)

## Discussion

This study evaluated the diagnostic accuracy, reasoning quality, and inference efficiency of four LLMs—ChatGPT-o3-mini, ChatGPT-o3-mini-high, ChatGPT-o1, and DeepSeek-R1—across 30 rare and complex disease cases. Diagnostic accuracy ranged from 30.0% to 40.0%, with no significant differences between models (Cochran’s Q test, P = 0.162), consistent with prior studies.^49,50^ ChatGPT-o1 demonstrated superior reasoning quality (4.08 ± 0.82), likely due to its advanced CoT optimization^51^, outperforming other models (P < 0.05). ChatGPT-o3-mini-high showed similar diagnostic accuracy (36.7%) to ChatGPT-o3-mini (36.7%) and slightly lower reasoning scores (3.69 ± 0.80 vs. 3.71 ± 0.90; P > 0.05), suggesting that its enhancements do not significantly improve performance in this context. Notably, ChatGPT-o3-mini-high did not demonstrate superior reasoning quality compared to its base model (o3-mini), despite advertised enhancements.^52^

DeepSeek-R1’s reasoning performance was better with English inputs (3.86 ± 0.86) than Chinese inputs (3.67 ± 0.84), despite its development for Chinese-speaking users. ^53^ This may reflect a predominance of English-centric training data or differences in language structure affecting reasoning clarity. Inference times varied significantly, with ChatGPT-o3-mini being fastest (7.0 ± 3.8 s) and DeepSeek-R1 (English) slowest (46.5 ± 32.5 s). The longer inference times for DeepSeek-R1 may be due to its mixture-of-experts architecture, which may involve complex routing and output generation.

Inference times varied significantly across models (one-way ANOVA, p < 0.001). ChatGPT-o3-mini was fastest (7.0 ± 3.8 s; range 3–17 s), while DeepSeek-R1 required longer times for English inputs (46.5 ± 32.5 s; range 16–161 s) than Chinese inputs (30.7 ± 22.7 s; range 11–137 s). This discrepancy likely stems from DeepSeek-R1’s mixture-of-experts (MoE) architecture,^50^ which increases token counts and computational complexity in CoT decoding, particularly for English prompts.

During inference, we observed two operational issues explaining model performance differences. DeepSeek-R1 intermittently returned ‘Server Busy’ errors, whereas ChatGPT remained consistently available, which may reflect backend resource constraints on the DeepSeek platform. Notably, when such errors occurred, DeepSeek-R1 failed to generate outputs. These instances were excluded from reasoning evaluation but did not affect the recorded inference time, as timing was measured only for successful completions. Additionally, under complex prompts, DeepSeek-R1 often produced repetitive or circular reasoning loops—potentially reflecting a reinforcement-learning reward scheme that prioritizes final outcomes over rigorous CoT reasoning.

The evaluation methodology employed in this study combined clinical expertise with advanced AI assistance. The hybrid evaluation panel, which consisted of two board-certified physicians from distinct specialties and one large language model evaluator (ChatGPT-4o), facilitated integration of human judgment with AI-driven assessment. This approach enhanced objectivity and reproducibility while maintaining essential clinical standards. A rigorous double-blind evaluation—where model outputs were anonymized and presented in random order—minimized recognition bias. The structured discrepancy resolution protocol, involving calibrated discussions and third-party adjudication, further reinforced scoring consistency. Inter-evaluator reliability was high (ICC = 0.84; 95% CI, 0.80–0.88), substantiating the framework’s robustness. However, inclusion of an LLM evaluator may introduce systematic biases due to model-specific preferences or errors. The limited size of the physician panel may also constrain generalizability, suggesting future studies expand human evaluator participation and include additional calibration rounds to bolster reliability.

### Limitations and Future Directions

Several limitations of this study should be noted. First, our analysis used a relatively small sample of 30 cases. Though diverse, this limits statistical power and external validity. Future work should incorporate larger, more heterogeneous datasets to enhance generalizability. Second, inputs were limited to standardized textual descriptions, which does not leverage the full multimodal capabilities of modern LLMs. More comprehensive assessments could be achieved by integrating direct analysis of medical images or physiological waveforms. Third, the use of an LLM as an evaluator, while innovative, is a relatively unvalidated approach that warrants further investigation. Despite high inter-evaluator agreement (ICC = 0.84), future studies should benchmark this AI-assisted framework against broader panels of human experts. Fourth, while our methodology enhanced objectivity, we did not investigate potential biases related to the reasoning style (e.g., verbose vs. concise) of the LLM outputs and their impact on evaluators.

Future research should address these limitations by assessing the real-world utility of these models through prospective clinical trials, for example, by integrating AI-generated differential diagnoses into clinical workflows. Additionally, performance may be enhanced through domain-specific fine-tuning or the use of retrieval-augmented generation. Finally, to optimize usability, user interfaces should be designed with cognitive load theory in mind, perhaps offering a dual-mode presentation with both rapid answers and detailed reasoning traces.

## Conclusion

This study demonstrates that advanced LLMs exhibit measurable capabilities in diagnosing rare and complex diseases. Their transparent reasoning pathways offer valuable clinical insights, fostering physician trust essential for real-world adoption. ChatGPT-o1 showed slightly stronger overall performance, but its proprietary nature limits on-premises deployment. Conversely, DeepSeek-R1’s open-source framework enables local customization and enhanced data control. Selecting an AI model for clinical practice requires balancing diagnostic accuracy, reasoning transparency, data-privacy compliance, inference latency, operational costs, and deployment feasibility.

## Data Availability

All data produced in the present study are available upon reasonable request to the authors

